# Peripheral capillary rarefaction is associated with cerebral small vessel disease burden: a population-based study

**DOI:** 10.64898/2026.05.05.26352496

**Authors:** Oscar H. Del Brutto, Denisse A. Rumbea, Robertino M. Mera, Fernando Góngora-Rivera, Eduardo J. Guzmán, Carlos Ríos, Emilio E. Arias, Víctor J. Del Brutto

## Abstract

**Background:** Peripheral microvascular abnormalities may reflect systemic microvascular dysfunction relevant to cerebral small vessel disease (cSVD), yet their relationship to individual neuroimaging markers and overall cSVD burden remains unclear. We evaluated whether abnormalities in nailfold capillaroscopy (NFC) are associated with specific cSVD markers and with the total cSVD score in a population-based cohort.

**Methods:** Atahualpa residents aged ≥60 years underwent NFC and brain MRI. Capillary tortuosities, dilatations, density, and megacapillaries were quantified using automated software with expert validation. Neuroimaging markers included white matter hyperintensities (WMH), lacunes, deep cerebral microbleeds (CMB), and enlarged basal ganglia perivascular spaces (BG-PVS). Logistic regression models assessed associations between NFC abnormalities and cSVD markers. Poisson regression was used to model the total cSVD score. All models were adjusted for demographics, educational attainment, and cardiovascular risk factors.

**Results:** Among 289 participants (mean age 71.3 ± 7.5 years; 51% women), lower capillary density was independently associated with CMB (OR: 0.70; 95% C.I.: 0.51–0.96) and lacunes (OR: 0.67; 95% C.I.: 0.50–0.91), with a borderline association for WMH (*p*=0.062). Megacapillaries were independently associated with moderate-to-severe WMH (OR: 5.01; 95% C.I.: 1.42–17.68). Tortuosities and dilatations showed no significant associations. Higher capillary density was inversely associated with the total cSVD score (β: -0.179; 95% C.I.: -0.283 to -0.075).

**Conclusions:** Reduced capillary density and megacapillaries track with the burden of cSVD. NFC may provide a noninvasive window into cerebral microvascular health and could inform risk stratification for cSVD progression and related outcomes.

## INTRODUCTION

Cerebral small vessel disease (cSVD) encompasses a range of microvascular processes that manifest through several neuroimaging markers, in particular white matter hyperintensities (WMH), lacunes of vascular origin, cerebral microbleeds (CMB), and enlarged basal ganglia perivascular spaces (BG-PVS).^1^ Although these markers are often combined into a total cSVD score,^2^ they differ in their pathophysiological specificity and likely reflect heterogeneous mechanisms beyond pure small-vessel pathology.

Peripheral microvascular assessment provides a window into systemic microvascular phenotypes that may mirror cerebrovascular injury. However, whether peripheral and cerebral microvascular abnormalities align remains largely unexplored, and it is still unclear which peripheral alterations best capture the burden of cSVD or its individual neuroimaging markers.

In a recent study, we reported that the presence of megacapillaries on nailfold capillaroscopy (NFC) was associated with greater WMH burden, suggesting that selected peripheral abnormalities may mirror cerebral microvascular damage.^3^ However, that work focused on a single NFC feature and a single neuroimaging marker, leaving open whether a broader peripheral microvascular phenotype relates to the overall burden and distinct components of cSVD.

Whether peripheral capillary abnormalities represent a systemic microvascular signature aligned with neuroimaging markers of cSVD remains unknown. We therefore aimed to evaluate the association between NFC abnormalities, capturing complementary dimensions of peripheral microvascular dysfunction,^4^ and both the presence and severity of individual cSVD markers, as well as the total cSVD score. This approach enables a more comprehensive assessment of whether specific or cumulative peripheral microvascular alterations preferentially correspond to neuroimaging markers that more directly reflect underlying cerebral small vessel pathology.

## METHODS

### Data availability statement

Aggregated data from this study are available from the corresponding author upon reasonable request.

### Study Population

This study was conducted in Atahualpa, a rural Ecuadorian village where several population-based investigations on cSVD have been carried out.^5–7^ Residents share relatively homogeneous demographic, socioeconomic, and lifestyle characteristics, including similar ancestry, educational attainment, dietary patterns, and limited access to healthcare.^8^ This uniformity reduces confounding from environmental and social variability and strengthens internal validity.

### Study Design

All adults aged ≥60 years who remained active participants in the Atahualpa Project cohort during the most recent survey (2025) were invited to participate. The cohort was originally assembled through annual door-to-door surveys initiated in 2012, with ongoing follow-up to document incident clinical events and vital status. To minimize alternative causes of microvascular injury, individuals with clinical, laboratory, or neuroimaging evidence of hereditary forms of cSVD, demyelinating disorders, or systemic autoimmune conditions known to affect the microcirculation were excluded.

After providing written informed consent, participants underwent updated cardiovascular risk assessments, brain MRI, and NFC on the same day. The primary objective of the present study was to evaluate the association between NFC abnormalities and both the total cSVD score and its individual components. All procedures adhered to the ethical principles of the Declaration of Helsinki. The study was approved by the ethics committee of Hospital-Clínica Kennedy in Guayaquil, Ecuador (FWA 00030727).

### Nailfold Capillaroscopy

NFC was used to quantify peripheral microvascular abnormalities following standardized protocols.^9,10^ Examinations were performed with an Optilia Digital Capillaroscopy System (Optilia Instruments AB, Sweden) equipped with ×200 magnification and integrated LED illumination. Images from the 2^nd^ to 5^th^ fingers of both hands were acquired and processed using Capillary.io (Capillary Technology SL, Spain). This automated software detects and classifies the proportion of tortuosities, enlarged capillaries (loops 20-50μm in diameter), megacapillaries (loops >50μm, often with distorted morphology), and capillary density (per mm) (Figure 1). Microhemorrhages – another frequent NFC abnormality – were excluded because, in this predominantly manual-labor population, the high frequency of fingertip trauma increases the likelihood of mechanically induced nailfold hemorrhages, reducing their specificity as markers of microvascular injury. All studies were performed by a single trained operator and reviewed by an expert reader blinded to clinical and neuroimaging data. Visual interpretations were used to confirm proper acquisition, support the automated outputs, and ensure consistency. Participants with low quality exams or discordance between automated and manual readings, underwent repeated NFC by the rheumatologist expert.

**Figure 1.**
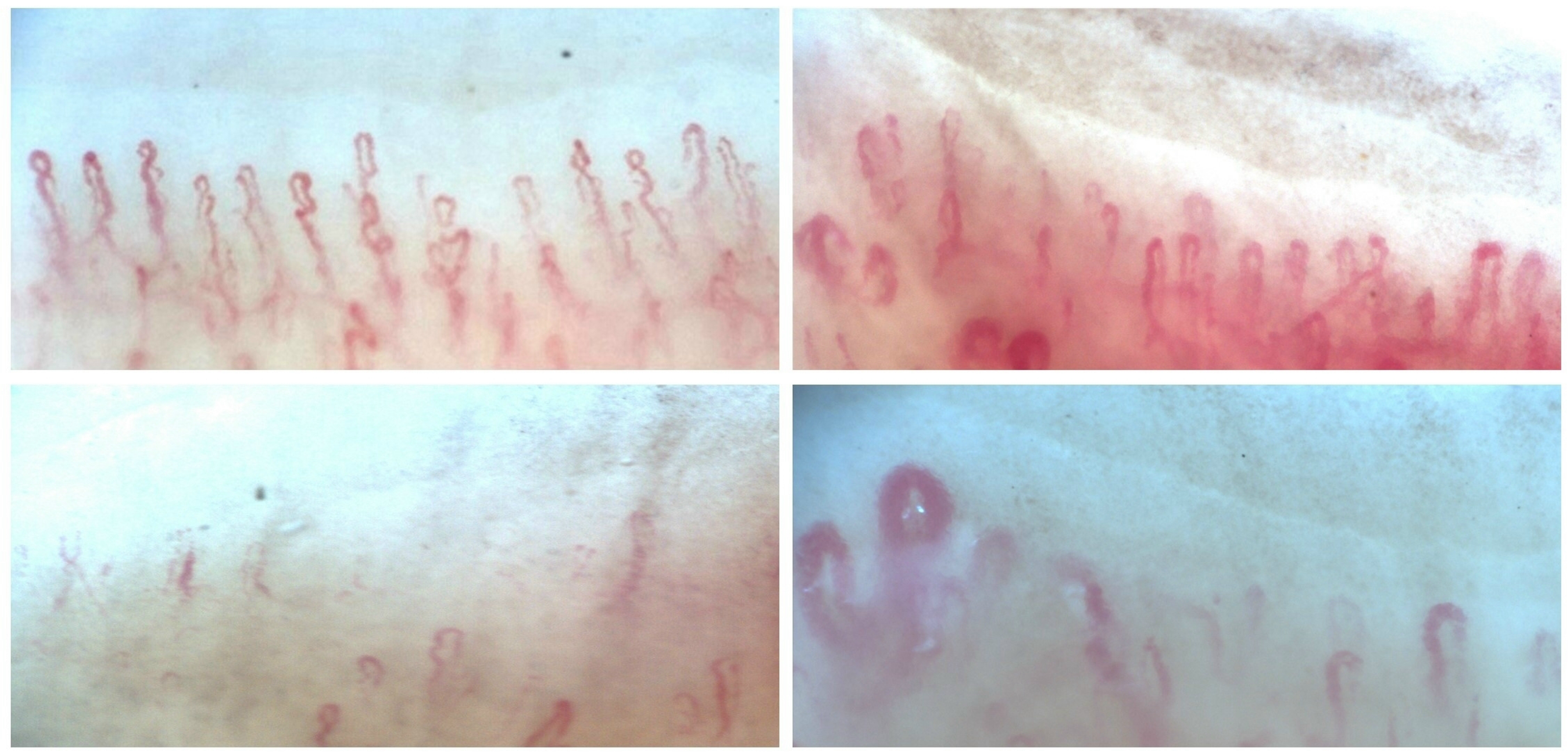
Characteristic abnormalities identified by nailfold capillaroscopy, including tortuosities (left upper panel), dilated capillaries (right upper panel), reduced capillary density (left lower panel), and megacapillaries (right lower panel).

### Neuroimaging Protocol

Brain MRI was performed on a 3T SIGNA™ Pioneer system (GE Healthcare, USA), using a standardized protocol aligned with STRIVE-2 recommendations for research on cSVD.^1^ The imaging protocol included axial and coronal FLAIR and T2-weighted sequences, a high-resolution 3D T1-weighted sequence, and a susceptibility-weighted angiography (SWAN) sequence. All exams followed the manufacturer’s predefined brain imaging package to ensure consistency across examinations.

Neuroimaging evaluation focused on the four conventional markers of cSVD: WMH, lacunes of vascular origin, deep CMB, and enlarged BG-PVS. Strictly lobar CMB were not considered because many of these lesions are attributable to cerebral amyloid angiopathy rather than cSVD. Because calcified parenchymal cysticerci are common in this population and may mimic CMB on susceptibility-based sequences, all participants also underwent non-contrast head CT using a standardized protocol on a Philips Brilliance 64 scanner. CT findings were used to differentiate true CMB from calcified cysticerci, thereby improving diagnostic specificity.^11^

WMH were defined as hyperintense lesions on T2-weighted images that remained bright on FLAIR without cavitation and were graded using the modified Fazekas scale.^12^ Lacunes of vascular origin were identified as round or ovoid fluid-filled cavities 3–15 mm in diameter located in the territory of a perforating arteriole, confirmed on T1-weighted images.^13^ Deep CMB were rated on SWAN using the Microbleed Anatomical Rating Scale, which requires the presence of well-defined, small, rounded hypointense lesions in deep brain structures.^14^ Enlarged BG-PVS were defined as <3mm linear or ovoid CSF-intensity structures following the course of perforating arteries.^15^ To derive the total cSVD score, one point was assigned for each of the following: moderate-to-severe WMH, at least one deep CMB, at least one lacune, and >10 BG-PVS in a single basal ganglia slice on one side of the brain (Figure 2), yielding a score ranging from 0 to 4.^2^

**Figure 2.**
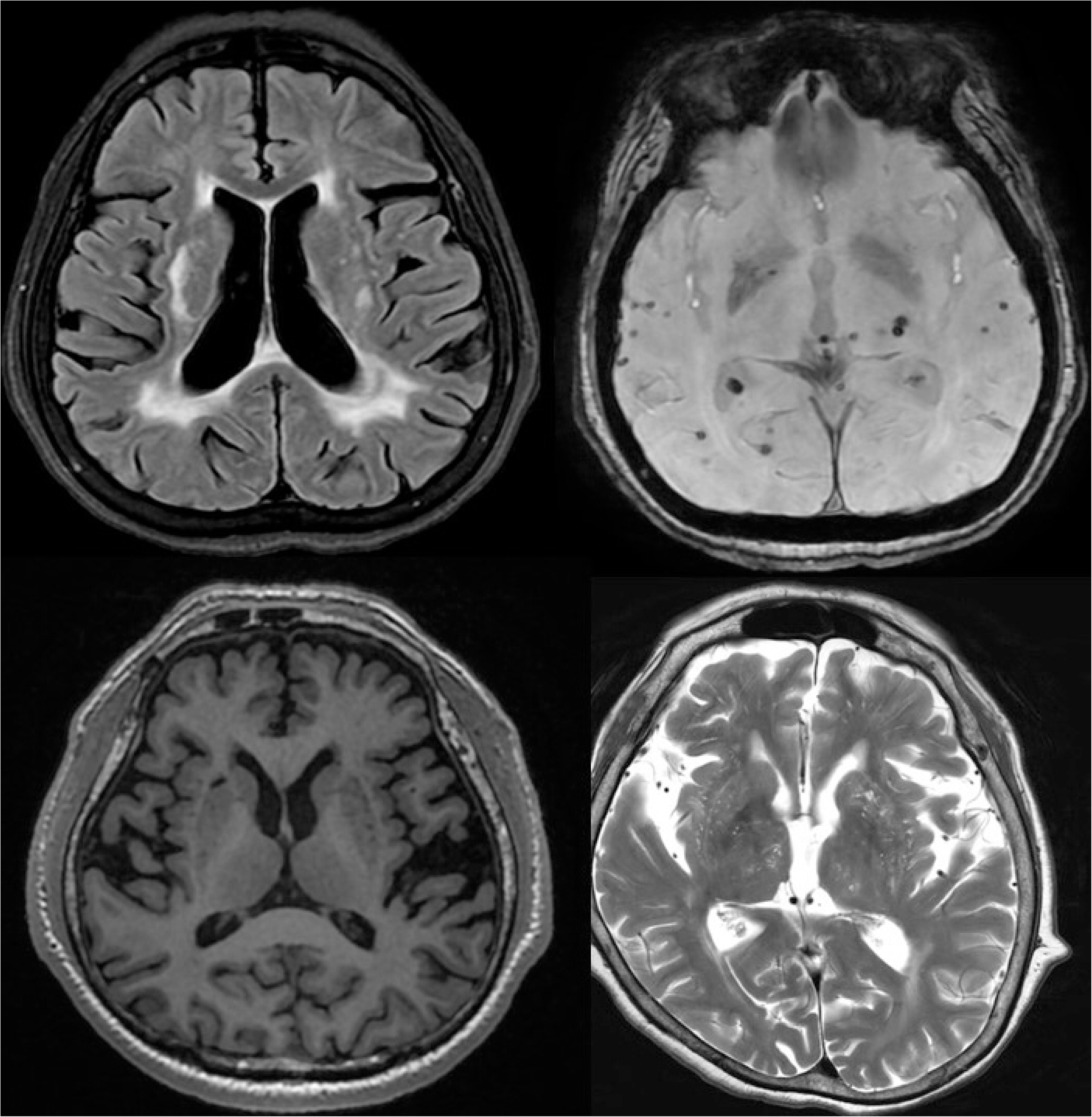
Neuroimaging markers of cerebral small vessel disease contributing to the total cSVD score: moderate-to-severe white matter hyperintensities (left upper panel), deep cerebral microbleeds (right upper panel), lacunes of vascular origin (left lower panel), and abnormally enlarged basal ganglia perivascular spaces (right lower panel).

All scans were independently reviewed by two experienced raters blinded to clinical and NFC data. Interrater agreements were from very good to excellent (κ=0.81–0.93) across all cSVD markers, and discrepancies were resolved by consensus with input from a senior neuroradiologist.

### Clinical Covariates

Demographic characteristics, educational attainment, and cardiovascular health metrics were included as covariates because of their established relevance to cSVD and peripheral microvascular injury. Cardiovascular risk factors were operationalized according to the American Heart Association’s Life’s Essential 8 framework,^16^ which evaluates eight domains of cardiovascular health: diet quality (assessed using the DASH score), physical activity, nicotine exposure, sleep duration, body mass index, blood lipids, blood glucose (fasting glucose and HbA1c), and blood pressure. Each metric was categorized following recommended thresholds to ensure comparability with prior population-based studies.^16^

### Statistical Analysis

All analyses were performed using STATA version 19 (College Station, TX, USA). Continuous variables were summarized using means and standard deviations or medians and interquartile ranges, as appropriate, and compared across groups using linear models. Categorical variables were compared using chi-square or Fisher’s exact tests. Primary analyses evaluated the association between each NFC biomarker and each individual cSVD neuroimaging marker using unadjusted and multivariate logistic regression models. Because the cSVD score reflects counting process of increased microvascular injury, we also fitted a multivariate Poisson regression model to estimate the association between each NFC abnormality and the increasing total cSVD score. Model diagnostics included assessments of multicollinearity, goodness-of-fit, and influential observations. Variance inflation factors were examined for all NFC variables, and no evidence of multicollinearity was detected. All multivariate models were adjusted for demographics, educational attainment, and cardiovascular health metrics.

## RESULTS

Of 360 community-dwelling adults aged ≥60 years enrolled in the Atahualpa Project cohort, 49 fulfilled at least one of the predefined exclusion criteria. An additional 12 individuals were unable to undergo MRI because their level of disability precluded safe transportation, five were ineligible due to claustrophobia or the presence of pacemaker devices, and five had non-interpretable NFC recordings owing to poor visibility of capillaries. The final sample included 289 participants.

The mean age of the study population was 71.3 ± 7.5 years (median, 71 years); 147 (51%) were women, and 184 (64%) had attained only elementary-level education. According to the Life’s Essential 8 framework, the proportions of individuals classified as having poor status in each health metric were as follows: 1) suboptimal diet, defined as a DASH score below the 25th percentile (n = 133; 46%); 2) low physical activity, with ≤29 minutes of moderate-to-vigorous activity per week (n = 7; 2%); 3) current tobacco use, recent cessation (<1 year), or use of inhaled nicotine delivery systems (n = 8; 3%); 4) inadequate sleep duration (n = 68; 24%); 5) obesity, defined as a body mass index ≥30 kg/m² (n = 74; 26%); 6) elevated non-HDL cholesterol (≥160 mg/dL; n = 98; 34%); 7) impaired glucose metric (<40 points; n = 69; 24%); and 8) uncontrolled blood pressure, with systolic values ≥140 mmHg or diastolic values ≥90 mmHg (n = 113; 39%).

The mean percentage of tortuous capillaries was 8.2 ± 5% (median: 7.4%) and that of dilated capillaries was 10.1 ± 10.0% (median: 7.8%). The crude mean capillary density was 10.5 ± 1.3 x mm (median: 10.6 x mm). Megacapillaries were observed in 15 participants (5%). On MRI, 96 individuals (33%) had moderate-to severe WMH, deep CMB were present in 34 (12%), lacunes in 40 (14%), and abnormally enlarged BG-PVS in 88 (30%) subjects. The total cSVD score was 0 in 152 individuals (53%), 1 in 62 (21%), 2 in 43 (15%), and 3–4 in 32 (11%).

Differences in demographic and clinical characteristics across NFC findings are summarized in Supplementary File 1. Briefly, participants with greater capillary tortuosity were more often men, whereas no significant differences were observed across categories of capillary dilatations. Higher capillary density was more frequent among women, and no additional demographic and clinical gradients were identified. The presence of megacapillaries was uncommon and showed no clear association with participants’ characteristics.

Before modeling the main variables, we performed crude group comparisons to explore unadjusted differences in NFC abnormalities across cSVD markers. Capillary density was lower in individuals with CMB (10.1 ± 1.5 vs. 10.6 ± 1.3; *p*=0.040) and lacunes (10.1 ± 1.7 vs. 10.6 ± 1.2; *p*=0.023) and showed a decreasing gradient across categories of the total cSVD score (*p*=0.022). Megacapillaries were more frequent among participants with moderate-to-severe WMH (10.4% vs. 2.6%, *p*<0.001). No significant crude differences were observed for tortuosities or dilatations.

Unadjusted models containing all NFC variables yielded results consistent with the crude comparisons (data not shown). We then fitted multivariate regression models to evaluate the independent association between NFC abnormalities and cSVD markers. Logistic regression was used for individual MRI markers (binary outcomes). After adjustment for demographics, educational attainment, and cardiovascular health metrics, lower capillary density remained associated with both CMB (OR: 0.70; 95% C.I.: 0.51 – 0.96) and lacunes (OR: 0.67; 95% C.I.: 0.50 – 0.91). A borderline association was observed between reduced capillary density and moderate to severe WMH (*p*=0.062). No independent associations were identified for tortuosities or dilatations and any cSVD marker. In addition, the presence of megacapillaries was independently associated with moderate-to-severe WMH (OR: 5.01; 95% C.I.: 1.42 – 17.68), with no significant relationship observed for CMB, lacunes, or enlarged BG-PVS. As expected, older age remained independently associated with all four neuroimaging markers of cSVD, while high blood pressure was associated with WMH and lacunes, and obesity with lacunes. For visualization, Figure 3 presents forest plots derived from the logistic regression models.

**Figure 3.**
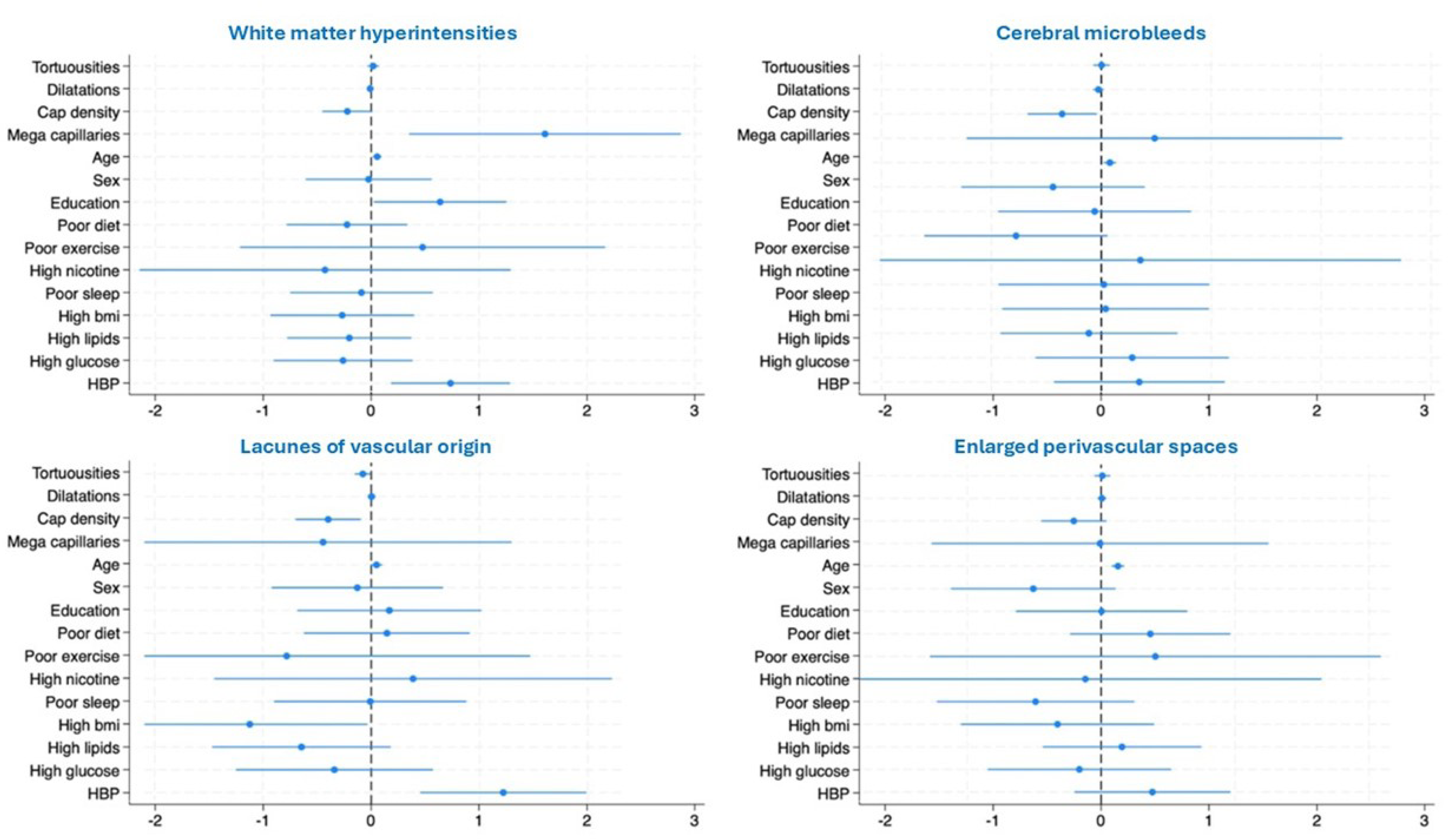
Forests plots showing associations between the different neuroimaging markers of cerebral small vessel disease and nailfold capillary abnormalities.

Additionally, we fitted a Poisson multivariate model to evaluate the independent association between NFC abnormalities and the total cSVD score. The analysis showed that higher capillary density was independently associated with a lower cSVD score (β: -0.179; 95% C.I.: -0.283 to -0.075), whereas tortuosities, dilatations, and megacapillaries showed no significant associations (Table 1). The magnitude and direction of these associations were consistent with the crude comparisons and with the logistic models used for individual MRI markers.

**Table 1.** Multivariate Poisson regression model showing associations between nailfold capillary abnormalities and the total cerebral small vessel disease (cSVD) score.

| **Total cSVD score** | **$\beta$ coefficient** | **95% C.I.** | ***p* value** |
| --- | --- | --- | --- |
| Tortuosities | 0.001 | -0.023 to 0.026 | 0.902 |
| Dilated capillaries | -0.004 | -0.018 to 0.010 | 0.592 |
| Capillary density | -0.179 | -0.283 to -0.075 | 0.001* |
| Megacapillaries | 0.026 | -0.544 to 0.597 | 0.928 |
| Age, years | 0.048 | 0.031 to 0.065 | <0.001* |
| Women | -0.188 | -0.464 to 0.088 | 0.182 |
| Primary school | 0.179 | -0.126 to 0.483 | 0.250 |
| Poor diet | -0.080 | -0.342 to 0.183 | 0.552 |
| Poor physical activity | -0.003 | -0.743 to 0.737 | 0.994 |
| Nicotine exposure | -0.024 | -0.813 to 0.766 | 0.953 |
| Poor sleep | -0.127 | -0.445 to 0.192 | 0.437 |
| Obesity | -0.321 | -0.663 to 0.021 | 0.066 |
| High non-HDL cholesterol | -0.073 | -0.339 to 0.193 | 0.590 |
| Diabetes mellitus | -0.110 | -0.422 to 0.202 | 0.490 |
| Arterial hypertension | 0.405 | 0.149 to 0.661 | 0.002* |

\* Statistically significant result

## DISCUSSION

In this population-based study of older adults, we found that reduced capillary density and the presence of megacapillaries were significantly associated with neuroimaging markers of cSVD. These findings extend prior work,^3^ and contribute to the growing evidence that peripheral microvascular phenotypes may mirror microvascular injury in the brain.^17–19^ By simultaneously evaluating multiple NFC biomarkers and the full spectrum of cSVD neuroimaging markers, our study provides a more integrated view of systemic microvascular health and its relationship to cerebral small vessel pathology. This multimodal approach allows the identification of shared microvascular remodeling patterns that may not emerge when individual features are examined in isolation.

The most consistent finding across analyses was the inverse association between capillary density and cSVD burden. In unadjusted comparisons, individuals with CMB and lacunes exhibited significantly lower capillary density than those without these lesions, and a clear decreasing gradient was observed across categories of the total cSVD score. The reproducibility of this pattern in fully adjusted models reinforces the interpretation that capillary rarefaction represents a robust structural marker of systemic microvascular dysfunction.

Capillary density is considered one of the most robust NFC biomarkers because it integrates chronic structural changes rather than transient functional fluctuations. Reduced density has been linked to hypertension, diabetes, and endothelial dysfunction,^20^ all of which are established contributors to cSVD. The fact that capillary density correlated with multiple cSVD markers, rather than a single imaging phenotype, supports the notion that it may capture a global microvascular vulnerability rather than a mechanism specific to any one lesion type. This aligns with the conceptualization of cSVD as a diffuse arteriolosclerotic process affecting small vessels throughout the body.

Megacapillaries, although uncommon, showed a strong association with moderate-to-severe WMH. These structures represent a marked morphological abnormality characterized by enlarged loops and distorted architecture. Although classically linked to autoimmune diseases, they may also arise in the context of chronic endothelial stress and maladaptive microvascular remodeling.^21,22^ The selective association with WMH, rather than with CMB or lacunes, suggests that megacapillaries may reflect a peripheral phenotype aligned with mechanisms such as chronic hypoperfusion, blood–brain barrier dysfunction, and impaired vasoreactivity, which are central to the development of WMH.^23^ This observation raises the possibility that distinct NFC phenotypes correspond to specific pathophysiological mechanisms within the cSVD spectrum.

In contrast, tortuosities and dilated capillaries showed no significant association with any cSVD markers or with the total cSVD score. These features are common in aging populations and may reflect benign or compensatory microvascular changes rather than structural injury.^9^ Tortuosity, in particular, increases with age and may be influenced by mechanical factors such as repeated fingertip use, which is highly relevant in this manual-labor community. Dilated capillaries are also nonspecific and may represent early or reversible endothelial responses rather than fixed structural abnormalities. Their lack of association with cSVD neuroimaging markers underscores the importance of distinguishing peripheral biomarkers with pathophysiological specificity, such as capillary density and megacapillaries, from nonspecific morphological variants that may be influenced by mechanical trauma or transient functional factors.

The lack of association between enlarged BG-PVS and any NFC abnormality is noteworthy and likely reflects the limited specificity of this neuroimaging marker for microvascular injury. Although enlarged BG-PVS are included in the conventional cSVD score,^1,2^ accumulating evidence suggests that they may arise from mechanisms beyond small-vessel pathology, including alterations in glymphatic clearance and sleep-related disorders.^24^ In this context, the absence of a peripheral microvascular correlate may indicate that enlarged BG-PVS capture a biological pathway that is only partially linked to systemic microvascular remodeling.

Several methodological strengths enhance the validity of our findings. First, the homogeneity of this population reduces confounding from socio-environmental factors. Second, NFC and MRI were performed simultaneously using standardized protocols, minimizing temporal variability. Third, the use of CT to differentiate true CMB from calcified cysticerci is a major advantage in this setting, where this parasitic disease is endemic.^11^ The study also benefits from careful neuroimaging adjudication, with high inter-rater agreement and consensus resolution by a senior neuroradiologist. Similarly, NFC readings combined automated quantification with expert review, ensuring both reproducibility and accuracy.

Despite these strengths, some limitations warrant consideration. The cross-sectional design precludes causal inference; whether peripheral microvascular abnormalities precede, accompany, or follow cerebral microvascular injury remains unknown. Therefore, these associations should be interpreted as correlational rather than causal. The homogeneity of this rural population enhances internal validity but may limit generalizability to more diverse populations. Megacapillaries were uncommon and estimates involving this phenotype have wide confidence intervals and should be interpreted with caution. While the intentional exclusion of microhemorrhages was justified, this decision removes a potentially informative marker of microvascular fragility. Finally, although the Life’s Essential 8 framework provides a comprehensive assessment of traditional cardiovascular risk factors, we were unable to evaluate inflammatory or genetic biomarkers that may modulate the relationship between peripheral and cerebral microvascular injury.

In conclusion, our findings support the concept that selected NFC abnormalities, particularly reduced capillary density and megacapillaries, reflect systemic microvascular dysfunction that parallels the burden of cSVD. Longitudinal and mechanistic studies will be essential to determine whether NFC could serve as a noninvasive tool to phenotype systemic microvascular health and potentially identify individuals at higher risk for cSVD progression.

## Data Availability

Data will be shared from the corresponding author upon reasonable request

## Non-standard Abbreviations and Acronyms

BG-PVS: basal ganglia perivascular spaces
CMB: cerebral microbleeds
cSVD: cerebral small vessel disease
NFC: nailfold capillaroscopy
WMH: white matter hyperintensities

